# Implementation challenges for an optimised surgery blueprint schedule in a children’s hospital: an exploratory qualitative study using the CFIR framework

**DOI:** 10.1101/2024.10.03.24314775

**Authors:** Kelly Vos, Theresia van Essen, Erwin Ista, Lonneke Staals, Saba Hinrichs-Krapels

## Abstract

Capacity management and planning for operating theatres involves scheduling surgeries, preparing beds, and ensuring available workforce. Numerous mathematical models have been developed in the scientific literature to optimise these schedules to aid planning decisions, but such models are rarely assessed for applicability and implementation within a hospital’s existing planning system. We address this implementation research gap by identifying the barriers and facilitators for adopting a mathematically optimised surgery blueprint schedule within a children’s hospital, using the Consolidated Framework for Implementation Research (CFIR). Facilitators for implementation of the schedule included a strong motivation among staff to optimise schedules given resource constraints and work burdens, as well as positive reactions towards an objectively designed mathematical scheduling tool. Barriers for implementation included a resistance to change among some staff, and the need for more evidence about the new schedule’s benefits before implementation. We identified a strong culture of operational adjustments made to schedules, which is a challenge to any optimised and centralised scheduling tool. Overall, we found that CFIR was a helpful tool for identifying specific innovation adoption factors for a proposed surgery scheduling model.

## 1. INTRODUCTION

The future sustainability of healthcare systems is highly dependent on the availability of its resources. The World Health Organisation (WHO) categorises these resources as human resources (workforce such as nurses and clinicians), materials and supplies (including medical devices and equipment), and medicines and drugs (Papanicolas et al., 2022). Planning these resources is particularly important in the operating theatre (OT) setting, where surgeries need to be scheduled, beds prepared, surgical tools ready, and, importantly, the workforce be made available. Despite accounting for a large proportion of hospital admissions and cost (around 70% according to (Denton et al., 2007)), the daily operations of OT departments worldwide are not always optimised. At worst, there are cancellations that impair patient’s health, the pressurised environment can lead to medical errors, and inefficiencies are created due to suboptimal planning.

State-of-the-art academic studies that resolve these inefficiency problems are in operations research and management sciences, which are extensively used in healthcare research and draw on applied mathematics, engineering and decision sciences. Given the interdisciplinary nature of these fields, the literature base is published across various academic disciplines, prompting the development of a tailored online database (Hulshof et al., 2011) to ease classification and searching for solutions to health(care) management and planning models for practitioners and researchers. However, these models are rarely tested for applicability and integration into a hospital’s existing planning system, both from the technology infrastructure and stakeholder acceptability perspective, creating an implementation gap. Indeed, there has been continued criticism and debate within the disciplines of operations research and management science on the applicability of models for the real world (Carter & Busby, 2023; Royston, 2013) and the need to bring evidence and empirical evaluations of operations research interventions into real-world healthcare settings (Lamé et al., 2022). Following an extensive literature review on studies specifically on OT planning and scheduling (the application domain of our study), Cardoen et al. note how very little is known about the process of implementation, or factors that enhance implementation (Cardoen et al., 2010). They state: “It is somehow contradictory to see that in a domain as practical as operating room planning and scheduling, so little research seems to be effectively applied.”(Cardoen et al., 2010) As summarised by Visintin et al, the two factors that are often cited as hampering the implementation of models in healthcare practice are, firstly, the limited involvement of stakeholders in model development, and, second, the excessive complexity of models that may not be compatible with the needs of the organisation or perceived as inadequate to address their problems (Eldabi, 2009; Visintin et al., 2017). It has also been argued that part of the implementation of operations research work happens outside of academia (see, for example, (van Lent et al., 2012) and (Brailsford et al., 2016)), which is why it is not captured in academic publications. This leaves an opportunity for a scientific approach to understand how implementation works in practice, thereby creating an empirical evidence base to enable future theory development of how implementation of these planning and scheduling models can be successful in other organisations.

In this study, we directly address this implementation gap and challenge for operations research and management science models, specifically in the healthcare domain. Our study’s aim was to identify the barriers and facilitators for introducing and/or adopting a mathematically optimised surgery blueprint schedule within a children’s hospital.

## 2. METHODS

### Overarching approach and theoretical framework

‘Implementation’ can mean different things in different scientific and practice communities. In a review by Brailsford et al., operations research and management science models are classified into three categories according to the extent of implementation: suggested to client (i.e. developed without interaction with a client organisation), conceptualised (i.e. developed through interaction with a client organisation) or implemented (i.e. used by a client organisation) (Brailsford et al., 2016). We adopt a similar definition to the third category of implementation: actual use by a client organisation. A growing discipline for examining the implementation of interventions is implementation science, described as “the scientific study of methods to promote the systematic uptake of research findings and other evidence-based practice into routine practice and, hence, to improve the quality and effectiveness of health services.”(Eccles & Mittman, 2006). We used the updated Consolidated Framework for Implementation Research (CFIR), which is specifically used to investigate the governance, behavioural and technical factors for introducing interventions and/or innovations in complex settings (Damschroder, Reardon, Widerquist, et al., 2022). In our case, the ‘setting’ in which the innovation was to be introduced was an academic children’s hospital, and the ‘innovation’ to be introduced was the surgery blueprint schedule (explained below).

At this children’s hospital, 18 different surgical specialties use the same shared resources such as the OT and wards. A new proposed schedule had been previously developed^1^ at the request of the children’s hospital to provide patients and their parents with more detailed information on when a specified patient (child) would have their surgery, to inform their planning as a family. In current practice, it is difficult to provide this date, and often surgeries need to be cancelled because of bed unavailability. This new schedule provides a blueprint of when groups of different surgery types can be scheduled (thereby it is more detailed than a ‘master surgery schedule’ (van Oostrum et al., 2008), since actual surgery groups are scheduled). The surgery group are then scheduled in such a way that the OT utilization is maximized and the bed occupancy at the ward satisfies the available capacity. The innovative part of this prior research is therefore that the scheduled surgery groups are sequenced over the day to also deal with the bed availability at the day care wards as patients need to be discharged at the end of the day. The blueprint can then be used by the surgeons or planners to assign specific surgeries to each scheduled surgery group. As a result, the resulting schedule (or ‘innovation’ according to the CFIR framework) is therefore in between the ‘tactical’ and ‘operational’ planning stages of OT planning for the hospital (Figure 1).

**Figure 1.**
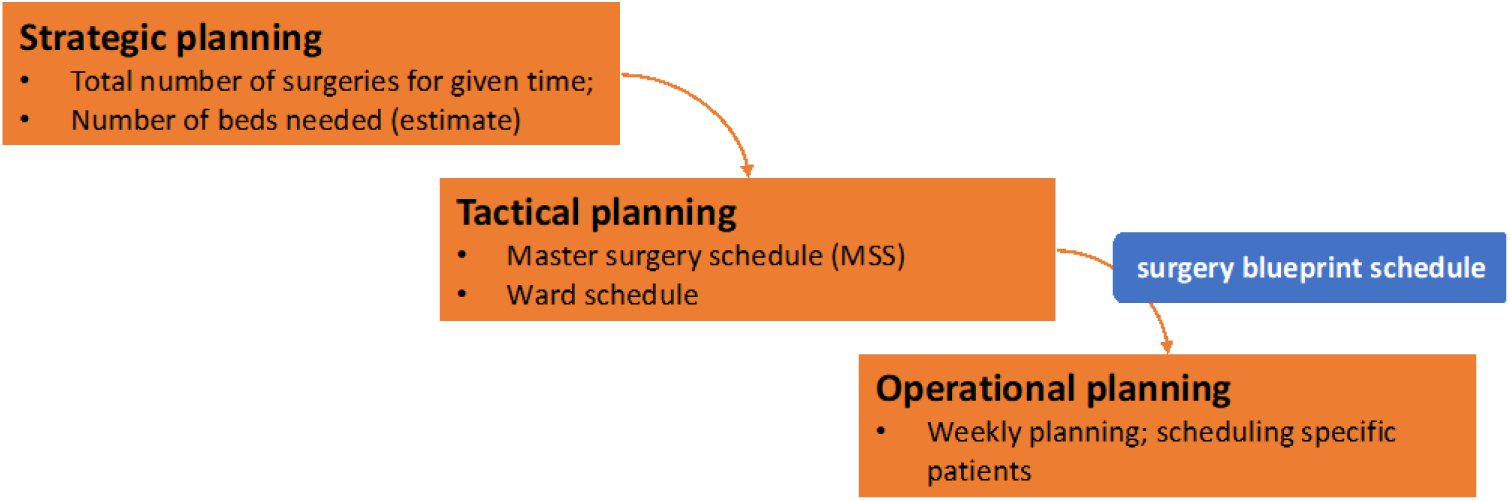
Different planning stages in the hospital

### Setting and participant selection

The setting for our study is the operating theatre and ward departments of a large (245 beds) academic children’s hospital in *[country redacted for review]*. For the purposes of our study, the planning department, operating theatres and wards in the children’s hospital were considered the ‘inner setting’ for the to-be adopted schedule (in accordance with CFIR guidelines). The wider hospital, and any further regulatory or policy drivers beyond the hospital were considered the ‘outer setting’.

In accordance with the CFIR guidance, we collected data from individuals who were seen to have power and/or influence over the implementation outcomes (Damschroder, Reardon, Opra Widerquist, et al., 2022). We initially used purposive sampling to identify participants. Participants were chosen for their involvement in scheduling and planning for beds, staff and/or materials in the operating theatres or wards at the children’s hospital. This included a variety of roles within the hospital with either exclusive capacity management responsibilities (e.g. planning coordinators, integral capacity management, admissions coordinators), as well as staff with clinical responsibilities who also are involved in planning (e.g. ward managers, nurses, surgeons, anaesthesiologists). These initial participants were identified by a discussion with representatives from the children’s hospital (one clinician and one capacity planning staff member); and subsequent participants were identified by referral from initial participants recruited for the study.

Staff were invited to participate in the study via email, as well as during a meeting for the planners in which the aims of the study were briefly presented. We estimate that a total of 25-30 staff were invited, given the number of people emailed, those attending the meeting, and those that would have been invited via word of mouth through staff who heard about the study. Of these, 19 were finally included in the sample (see Table 1 for full details of included participants in the study). Interviews were conducted face-to-face on site within hospital office spaces. Some interviews were held in groups, with only the interviewees and interviewer present.

**Table 1.** List of interview participants included in the study.

| Interviewee number* | Role in hospital | Role in planning and scheduling |
| --- | --- | --- |
| 1 | Admissions coordinator, Ward PICU | bed capacity PICU |
| 2a | Planning & Schedule coordinator (resources), OT | OT room, OT schedule, materials |
| 3a | Planning & Schedule coordinator (resources), OT | OT room, OT schedule, materials |
| 4a | Planning & Schedule coordinator (staff), OT | daily coordination OT |
| 5 | Sector manager (staff), OT | staff capacity OT/ member TPC |
| 6b | Admissions coordinator, Ward | bed capacity wards |
| 7b | Admissions coordinator, Ward | bed capacity wards |
| 8c | Manager, Daycare Ward | staff & bed capacity daycare ward |
| 9c | Senior nurse, Ward | bed capacity wards |
| 10 | Personnel/Team manager (staff), OT | staff capacity OT |
| 11 | Surgeon, Children (Planning Specialty Surgery only) | specialty specific patient planning/ member TPC |
| 12d | Anaesthesiologist, manager | daily coordination OT |
| 13d | Anaesthesiologist | member TPC / daily coordination OT |
| 14 | Surgeon, Children (Planning Specialty Surgery only) | specialty specific patient planning/ member TPC |
| 15 | Surgeon, Children (Planning Specialty Surgery only) | specialty specific patient planning / member OPC |
| 16e | Consultant integral capacity management | integral capacity management/ TPC |
| 17e | Consultant integral capacity management | integral capacity management/ TPC |
| 18 | Sector manager (resources), OT | OT room, OT schedule, materials |
| 19 | Planner (Planning specialty surgery only) | specialty specific patient planning / member OPC |

**Abbreviations:** Paediatric intensive care unit (PICU), Operating theatre (OT), Operational Planning Committee (OPC), Tactical Planning Committee (TPC)
\* All interviewees designated with a letter participated in a joint interview with another interviewee with the same letter

### Data collection

We conducted semi-structured interviews with the 19 participants. Interviews lasted circa 60 minutes and were conducted in *[language redacted for review]* as that was the language in which the participants and interviewee *[redacted for review]* spoke most fluently and comfortably. Interviews were recorded and transcribed verbatim. Transcripts were translated for internal analysis purposes, initially by one author *[redacted for review]* and fully checked by another author *[redacted for review]*.

The interview topic guide (Appendix A: interview guide) was designed by adapting the (updated) CFIR framework. Three researchers *[redacted for review]* consulted together and selected the questions most relevant to the intervention studied (the optimised surgery blueprint schedule), resulting in an adapted version of the CFIR questionnaire which we used for this study (see Appendix B, Table B1 for the use of each construct taken from the CFIR framework). In addition to the interview guide, participants were previously sent a flyer providing information about the surgery blueprint schedule about which they would be interviewed, shown in Figure 2 and Figure 3.

**Figure 2.**
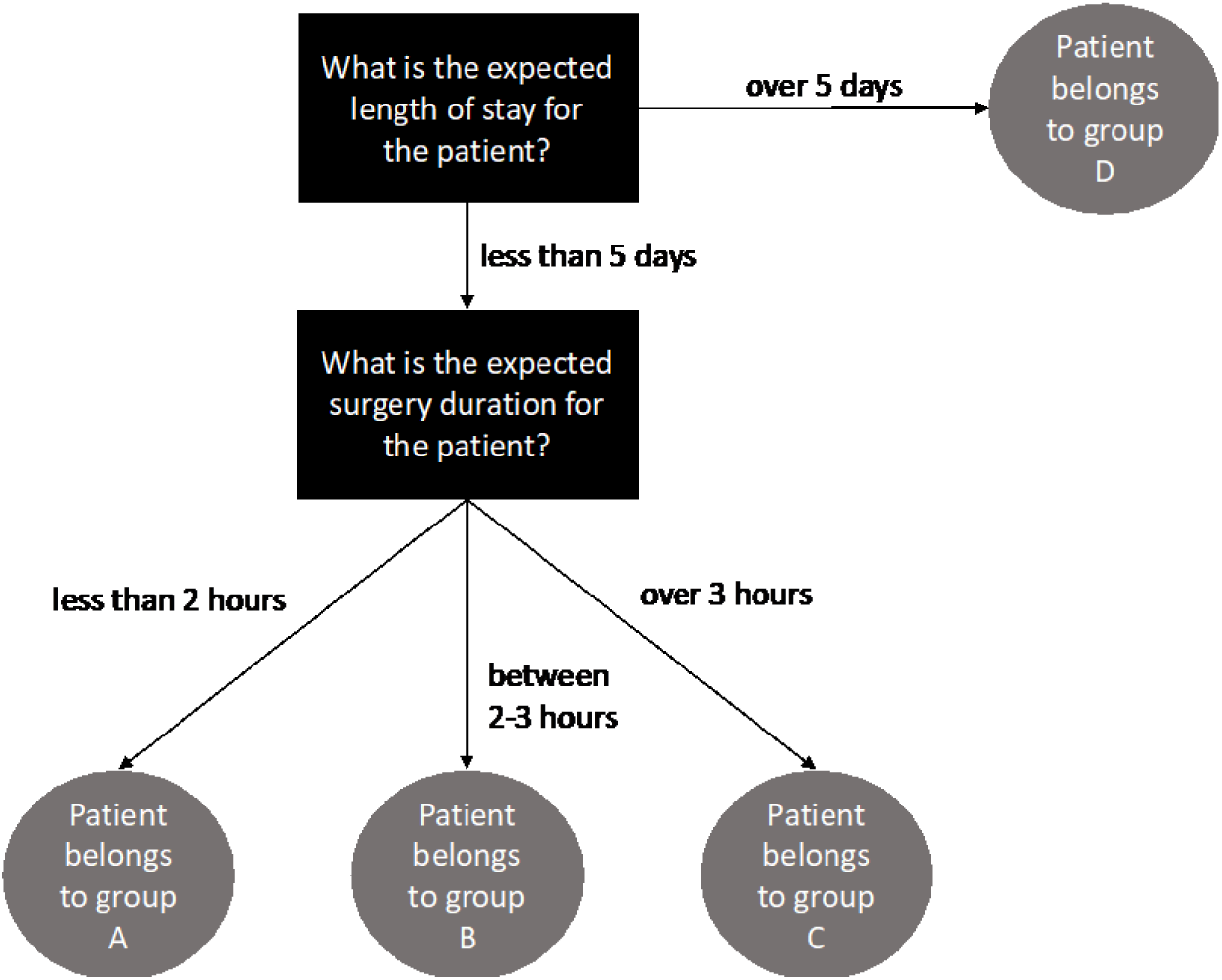
Sample of surgery blueprint schedule conceptualisation shown to show how patient groups are categorised according to surgery duration and length of stay

**Figure 3.**
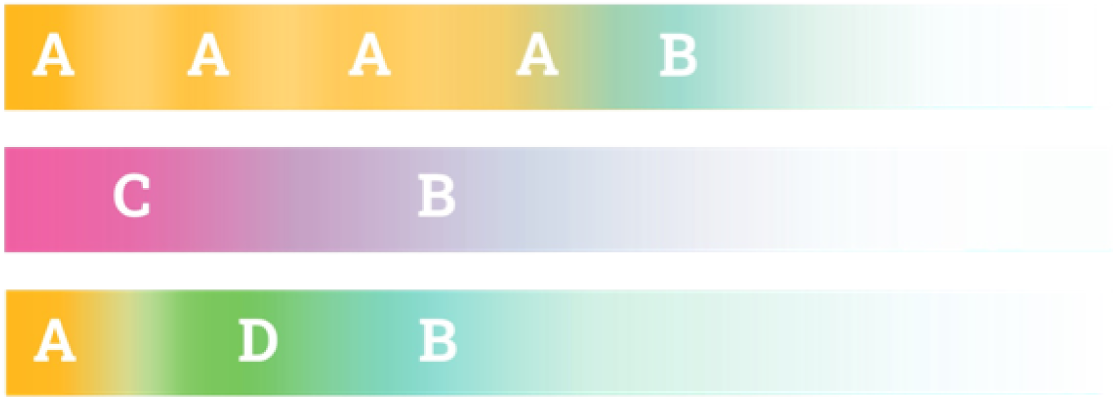
Surgery blueprint schedule conceptualisation (as presented to the interviewees). Each bar represents an operating room. The letters represent the scheduled surgery groups. The fading colours represent the uncertain surgery duration.

### Data analysis

Transcripts were uploaded onto ATAS.TI and analysed using thematic coding. Initial codes were sought and applied guided by the CFIR constructs (Appendix B), and further emergent codes were identified using open coding. One author coded the transcripts *[redacted for review]* and a selection of these were checked by another author *[redacted for review]*.

### Ethics statement

This study was approved by the Human Research Ethics Committee of the *[university name redacted for review]* considered a low-risk study (file number 3112). Informed consent forms were signed by all participants in the study.

#### Data sharing statement

Original audio recordings, transcripts and signed informed consent forms are kept on secure password protected servers (SURFDRIVE) available to the research team only, due to the inability to fully anonymise participants in the files. All other data available in this manuscript: interview protocol (Appendix A), selection of direct anonymised quotes from participants, and list of roles of participants (Table 1).

## RESULTS

### 3.1 Overview of interview participants and planning processes

A total of 19 staff participated in the interviews (see Table 1 for their characteristics and roles in relation to planning and scheduling). Given that some interviewees opted to participate an interview together with their colleague, 13 interviews were held.

These individuals play different roles within the planning process. An overview of these broad range of individuals involved in surgical planning, with varying responsibilities, is illustrated in Figure 4.

**Figure 4.**
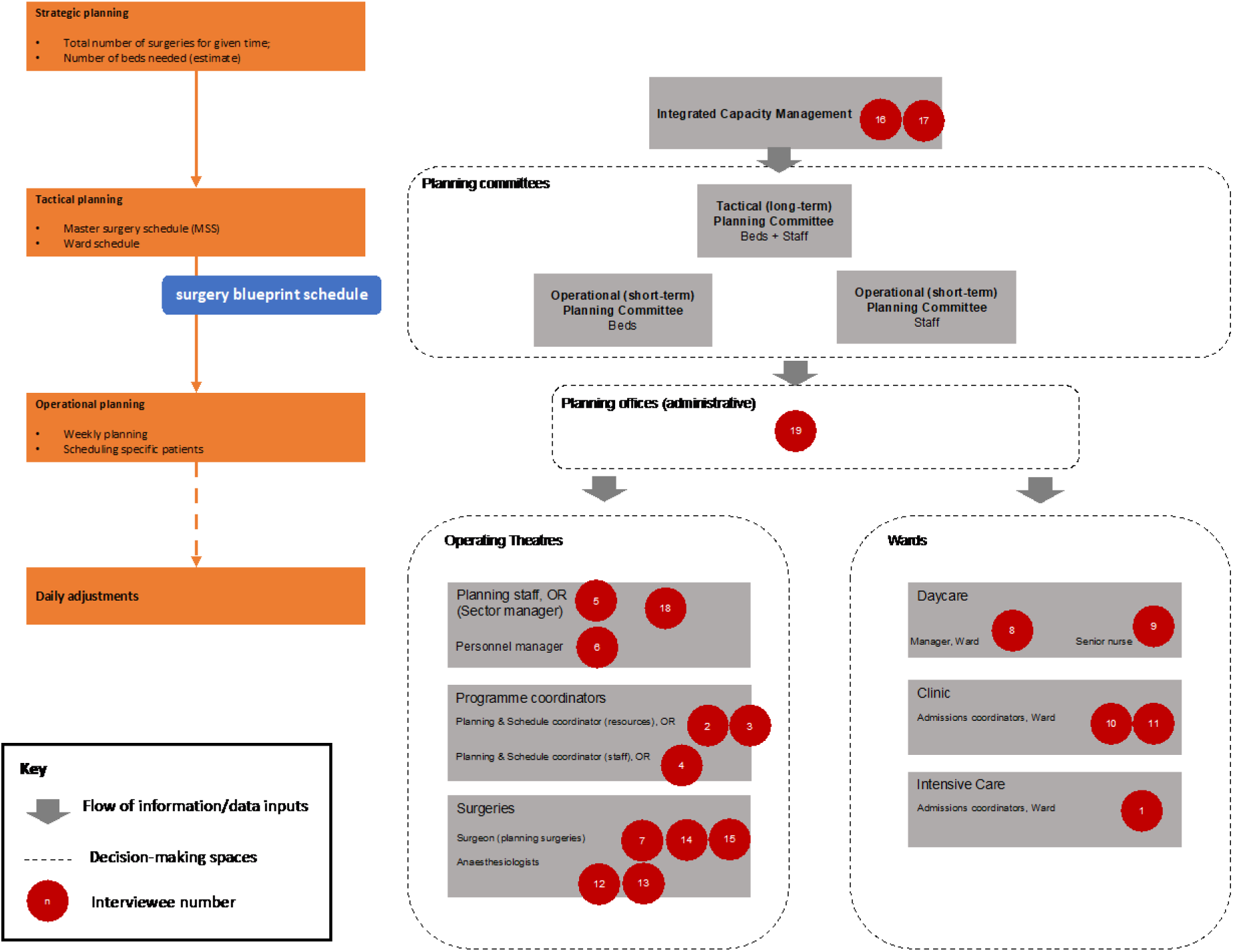
Overview of planning/scheduling staff interviewed, and relationship to tactical and operational planning functions (numbers correspond to interviewed participants in Table 1). Orange boxes demonstrate how these planning levels correspond with general planning in Figure 1. Blue box indicates the surgery blueprint schedule which is the ‘innovation’ introduced in this study.

Through our interviews we elicited how decision-making processes and information decision flows take place for the purposes of planning, as described briefly below (referring to Figure 4):

- **Strategic planning:** Master surgery schedules (blueprints) are originally created at this level, mostly on an annual basis. They provide a general structure for the total number of surgeries and number of beds needed, but these are annual estimates of total hours of use for the operating room per specialty.
- **Integrated capacity management:** This team has the oversight of the bed occupancy levels and staff numbers. This data is used as input for different planning committees (next bullet point).
- **Planning committees:** These committees plan how to fill beds and ensure there are available beds for the incoming patients. Tactical level planning (by the Tactical Planning Committee) is where ward schedules are planned. Master surgery schedules can be adapted slightly at this level by the committee. Operational level planning (by the Operational Planning committee) is where weekly schedules are created.
- **Surgery blueprint schedule:** This is the ‘innovation’ introduced in our study: it provides a blueprint of when groups of different surgery types can be scheduled (thereby it is more detailed than a ‘master surgery schedule’ created in the Strategic and Tactical planning phases, since actual surgery groups are scheduled. The blueprint can then be used by the surgeons or planners to assign specific patients to each scheduled surgery group.
- **Operational planning/Planning offices:** The daily filling of the blueprint schedule is completed at this level, by allocating actual patients (including contacting patients/families to see if they are available). In principles, this would be considered the final schedule.
- **Daily adjustments:** These are daily adjustments made to the allocation of patients, depending on staff and bed availability, and possible cancellations. This is done by the programme coordinators both in the operating theatres and in the wards.

We refer to these processes in the rest of our results. Of particular importance is to note that the proposed new blueprint schedule is designed to schedule ‘groups’ of patients only, according to their surgery type (including length of stay and expected surgery duration, see Figure 1). It does not allocate specific individual patients, as this is done further down (see Figure 4, under ‘planning offices’). We therefore refer to the new schedule in our results as a schedule for patient groups, and not a schedule for patients. We divide our results according to the constructs identified in the CFIR framework, as this guided both the interview protocol and the clustering of the initial themes in the analysis. We also report on additional insights on implementation which emerged during the analysis, beyond the constructs sought from our analytical framework. Constructs directly from CFIR are in **bold**.

### 3.2 Innovation domain

#### General positive reactions to a mathematically derived schedule

To make the proposed new way of patient group scheduling understandable, we created a sketch to demonstrate how it differed from the current master surgery schedule (see Figures 2 and 3), thereby communicating the intent of the proposed innovation. All interviewed participants could explain it back to the interviewer in their own words, suggesting they understood it.

All interviewed participants positively reacted to the proposed **innovation source**, which in this case was a mathematically derived schedule, describing it as “refreshing and innovative” [Interviewee 5], that it “makes a lot of sense” and is “good”. Participants also commented on how the model provides objectivity to planning.

> *“…taking the intuition out of it and doing it more on the numbers and say with evidence I think that’s better.” [Interviewee 11]*
>
> *“The maths is fair. Mathematics does not distinguish between my opinion and anyone else’s.” [Interviewee 14]*
>
> *“I think it’s really good to look at numbers, because otherwise I think you’re emotionally favoured there, that you think, “Well in that OT, only that is possible.” and that surgeon only wants to be there.” [Interviewee 10]*

Most participants pointed out the **relative advantage** of the new group scheduling compared to current practice. They acknowledged that such new group schedules are indeed needed, even if this proposed model was ‘not perfect’ nor taking into account the dynamic changes within the hospital environment.

> *“The biggest selling point is that it will make bed capacity more ‘even’ across the week (rather than peaks on Tuesday etc)” [Interviewee 7]*.
>
> *“Yes, because [for example] you have to cancel four children on Thursday and Friday, but on Monday and Tuesday there were eight places together.” [Interviewee 6]*

However, given that this was not a fully validated model/schedule, a few participants pointed out limitations of the schedule itself, for example that the duration of surgeries are not sufficiently taken into account. This was to be expected, given that this was not a fully validated and trialled innovation, but rather a proposed new schedule. Even for participants who were positive about the new schedule, they commented on the need for more ‘medical knowledge’ for it to be implementable, but they saw the potential in a more objective way of scheduling, and this was considered “refreshing” [Interviewee 5 and Interviewee 12].

> *“I’m really glad it’s being looked at this way because I think there’s really room for improvement [in current practice]” [Interviewee 11]*

Another pointed out how much of the current practice involves adjustments made while filling in patients within the current schedules. The new mathematical scheduling approach allows these rearrangements to happen earlier:

> *“We do this already, in terms of puzzling with the variables… but the difference with such a model is that it can do it for you, saves time in that puzzling.” [Interviewee 14]*

#### The required balance between centrally optimised group schedules and on-the-floor needs

Given that this was a proposed first version of a new patient grouping schedule, one of the main discussion points in the interviews was its ability to consider the complexity and dynamic behaviour of the operating theatre environment. Unprompted, a handful of respondents focused on the exceptional cases when paediatric patients need more time for comforting before a surgery, therefore stating that these schedules would not work in practice. They emphasized that children are unpredictable and indicate that they are therefore afraid that theory and practice are different. The schedule itself was therefore not deemed to take into account the level of adaption and complexity in the system by some of the participants, as this level of **adaptability** was not obvious yet in the proposed schedule (although, in reality, this was not proposed to be a centralised individual patient schedule, but a blueprint for assigning groups of patients, see Figure 1).

A few participants were happy to think along with the design of the schedule and commented on how such adaptability can be achieved by specific parameters that could be incorporated into the mathematical model, including factors that are already considered such as the surgery duration, length of stay of different patients, and new factors, such as the ‘difficulty’ of the patient (such as the presence of multi-morbidities for the patients, whether the child needs extra psychological or emotional support before/after surgery) and the variable amount of time needed by each surgeon or anaesthesiologist to perform their tasks (*“you need to take the human diversity in the workforce into account” [Interviewee 5]).* These latter factors are more difficult to assess and can affect the surgery groups assigned, or indeed affect the daily allocation of individual patients. Although these factors were mentioned across the interviews, they were not quantified or specified in detail, indicating a degree of tacit knowledge about how patients are expected to be different. This adds to their perceived need of always needing to make changes on the floor, no matter what a new novel way of group scheduling may bring.

> ***“…****sometimes you just have to think in advance “we have to have some extra time with this one”, so maybe there has to be some kind of way that you can put an extra block in of “needs more time [in the model]” [Interviewee 12]*

The interviewer explained that such factors could be considered by creating ranges and parameters within the model that can be adjusted, and this resonated well with the participants.

Finally, participants were keen to ensure that the schedule considers the whole chain of events in the hospital *(“It’s a chain and a domino” [Interviewee 3]*). Surgery changes affect ward occupancy, and not every department will have the same needs (reflected, partly, also on the staff working there).

> ***“****The most important thing is just that when it comes to it, it’s not just about us, it’s going to affect the whole hospital.” [Interviewee 2]*

Again, this is exactly what the proposed group schedule intends to do in a future fully developed version. But the emphasising of these adaptability points demonstrated that, despite understanding the intent of the new blueprint schedule in theory, there were still some perceptions that it would be more rigid or centralised than intended to some of the interview participants.

### 3.3 Outer setting domain

#### External pressures support innovation

What stood out as an external driver for adopting this new schedule, in the context of their current experiences, was the relatively urgent need to release pressure from staff (e.g. “*the nurses are reaching their breaking point [Interviewee 8].* Dissatisfaction with current scheduling arrangements was voiced by all participants, and therefore a more objective form of scheduling was generally welcome. Furthermore, it had been noted that the hospital board has also made explicit the requirements to create efficiencies, as pressures on the workforce and shortages of staff contribute to existing challenges.

> ***“****Yes, the need is mainly that we are again confronted today that there is a continuous shortage of staff, so there is a shortage of admission beds. So everyone really feels the urgency to get the most out of the availabilities, we all do our best for that, but actually we are all just kind of yes well-meaning volunteers, say amateurs, actually and there is as much as possible open in the field of algorithms or systems that can really help us. That people really do see that something must be better everything that makes it better, just have to grab it with both hands, yes.” [Interviewee 12]*

From the participants’ perspective, there are no **regulations, laws or policies** prohibiting the potential introduction of the proposed schedule innovation as a barrier to the adoption of such an innovation.

### 3.4 Inner setting domain

#### Informal communications as both enabler and potential barrier to implementing new schedule

Generally, the participants acknowledged ongoing communication between the different planners and planning departments, especially during the week when cancellations occur or when more beds are needed somewhere.

> *“In that respect, we try to think along with each other, because we all want the best for the patient.” [Interviewee 1]*
>
> *“If they need something, they just ask, so to speak. They don’t hesitate to ask. At the [Operational Planning Committee], it is also said if a bed becomes available, then I want it and then I also write it down … like if there is space then it takes precedence first.” [Interviewee 9]*

As described in Section 3.1, general blueprint schedules are formed centrally, but allocation of patients are created in consultation between clinical staff and coordinators.

> *“Every morning we [the PICU admissions coordinators, three nurses actually] walk around the beds and see who’s lying there, so then we see if we have empty beds and for the kids that are there, what’s the plan with them? Some just stay put and some can go to another hospital or to another ward and some can go home.” [Interviewee 1]*

A few participants who have clinical and/or surgical roles (e.g. interviewees 11-15) refer to one-on-one conversations they have with planners and coordinators to influence their changes in how patients are allocated. For example, they discuss with their planners the waiting lists, which types of surgeries on it involve more complex planning (cooperation with other departments, or needing an ICU bed) and should be given priority and filling the gaps with more elective cases, which can be rescheduled more easily if needed.

While these informal relational connections between staff are helpful in adopting such a new innovation as the proposed schedule, those who design any new proposed group schedule would have to bear in mind that such informal meetings may result in compromising the intention of a centralised blueprint schedule if it were to be too rigid. Currently, our proposed innovation creates a general blueprint but with some restrictions (i.e. by patient groups, as seen in Figure 1), but the actual patient allocation is up to ground-level decision-making. So, while there is flexibility to some extent on actual patient allocation, the adjustments on how these patient groups are scheduled centrally may be cause for resistance (as described in the next section). Therefore, while such open communication enables adoption of any new innovation, in this case it might create a barrier to renewed planning approaches, because such on-the-ground changes is so embedded in the working culture. A few participants went as far as stating that these small conversations between surgeons and planners are the “real planning”, and planners “fiddle with [the schedule] until it fits” [Interviewee 12]. These habits and working practices could work against the adoption of a centralised scheduling of patient groups.

##### The potential for resistance to change in existing working cultures

As alluded to above, there was a general perception by the interviewed staff that there could be resistance to change because of (clinician) individual preferences on their scheduling choices. That is to say, that even the way the patient group schedules are optimised (to take into account more efficient practices across the whole hospital) may be cause for resistance for some clinical staff who have individual preferences on where their surgery groups should be scheduled. Indeed, such resistance to change was an often-mentioned topic in the interviews. Taking away this autonomy and “freedom” (Interviewee 16) may create more resistance to any new scheduling approach.

In order to be able to bring stakeholders/planners along, participants mentioned that it would be important to emphasise what this grouped schedule is trying to do: in other words, describe the intention to more efficiently level bed occupancy for the whole hospital (including wards), rather than only creating optimised schedules for the operating theatre. There were also suggestions of how to create change, by taking people along as the intervention is being introduced, and recognize that changes take time.

> *“I first let it sink in, because I had already thought of that or that seemed smart to me, but then it is useful if I inquire “What do you think?” …the people who were actually going to change places, agreed, and two did not, but they indicated “In the long run, we would like that, but not now.” [Interviewee 10]*
>
> *To make sure it’s done well, you have to take everyone with you, because otherwise you’ll all get people saying, yes, it’s decided but I’m not doing it anyway. I think there are a lot of captains on the ship, who set their own course, so the chance of success is only if you try to get everyone on board.” [Interviewee 1]*

However, it should be noted that despite many participants mentioning such resistance to change, this sentiment did not appear among those we interviewed, which represented a range of roles and responsibilities within the operating theatres and wards.

### 3.5 Individuals domain

#### A range of individuals have influence over scheduling

As was shown in Figure 4, we observed that the same individuals are involved in centralised planning committees (named Operational Planning Committee and Tactical Planning Committee in Figure 4), as on the ward. Further, those involved in planning, either in a centralised capacity or within the operating theatres or wards, have different degrees of influence in creating changes in schedules. Despite the seemingly top-down setup in the figure and above description, as was noted in the previous sections, many changes are made lower down daily, when actual patients are allocated.

Throughout the interviews, we identified individuals who would ultimately have to be involved in **leading and facilitating** the new schedule; these included the Tactical Planning Committee, the operating theatre managers, ward managers, as well as the ‘user council’ which includes representation from all the medical specialities and wards. However, the **innovation recipients** in this case are clinicians who ‘receive’ the new OT schedule which changes both the schedule itself and the form of scheduling, and they are both affected by centralised decisions and display power to change these decisions (schedules), due to their preferences.

> *“Surgeon X always sits down with Y, the planner. And then finally determine the order who goes where. I don’t know if all planners do that, but to that extent the surgeon will have a finger in the pie there too.” [Interviewee 2]*

According to some participants, this can be problematic – despite benefits to the end-user (the clinician ultimately planning and benefitting from the revised schedule), decisions on schedules (for surgery groups) made at one ward create knock-on effects at other wards and their respective schedules, which may not be optimal, creating further inefficiencies. This was exactly what the new planning tool is trying to resolve through its optimised master blueprint schedule. However, when prompted to respond on who would be the **leaders and facilitators** of a new scheduling intervention, many mentioned ‘planners’, referring to the more administrative centralised planning (i.e. those further up in Figure 4), rather than the end-user clinicians who may, in some instances, actually have more influence over individual patient schedules.

Finally, we learned the importance of the diversity of skill set in capacity management in general, which can have an influence over how new innovations are taken up. Some new innovations may come across as too technically challenging, and require in-house expertise for future adoption and implementation, and such expertise may not always be present in current practice (although this can vary across hospitals).

> *“…so that it is then made from certain mathematical backgrounds or technical business administration certain models and then at least for me and I think for most of them are abracadabra. The outcome is clear, but how do I get to where we can build that up? Who is going to pick this up? Who’s going to keep up? For that you actually need that expertise that I think may be lacking or at least not sufficiently present.” (Interviewee 16)*

## 3. DISCUSSION

As with any other innovation for clinical practice, a change in management or organisation comes with implementation challenges. Using the CFIR framework, we identified specific barriers and facilitators to implementing a proposed operation room blueprint schedule in a children’s hospital. We divide our discussion into what we learned about the implementation challenges of the schedule (innovation) itself, what we learned regarding design inputs for the innovation in the future, and what we learned about the use of CFIR for exploring implementation challenges in this context.

### What we learned about the challenges for implementation of the surgery blueprint schedule (‘the innovation’)

Overall, we observed general positive reactions to the mathematically derived schedule, across the participants. In principle, it was acknowledged that a centralised blueprint group scheduling tool would be able to balance the needs of the operating theatres and wards, creating overarching efficiencies for the hospital. Participants expressed a clear need for this innovation [the new schedule], expressed almost unanimously, due to the pressures on staff right now and inefficiencies in planning. This attitude will be a key facilitator for new centralised planning tools. There are also lines of open communication between centralised tactical planners and those further in the operational planning process. Although resistance to change in the organisation can be expected, it was acknowledged that genuine engagement in the design of the tool and introducing changes incrementally could facilitate the adoption of a new scheduling approach. A desire was expressed for specific stakeholders to be brought along while the innovation [the new schedule] is being developed and ensure they feel part of the process, a pattern common to many organisations trying to implement change (van Bruggen et al., 2019). Finally, the advantages of the scheduling tool should be clearly communicated, specifically its ability to bring efficiency improvements to the whole organisation, rather than an individual ward, unit, or operating theatre.

However, our analysis identified that despite the intention to adopt centralised schedules, there are cultural and work practice barriers to overcome. The identified open lines of communications are both enablers and barriers to implementing a centralised schedule. Schedules created at the tactical level are adapted and adjusted to suit individual’s needs – which might benefit the specific surgery speciality, for example, but have negative consequences in other wards. This behaviour is common in other capacity management instances and relate to examples of local adaptation and decentralised adjustments, explaining changes made on the ground from a centralised plan. Given that the proposed surgery blueprint schedule in this study sits between tactical and operational planning (Figure 1), some degree of tactical flexibility and operational adjustments are necessary to make it function. Similar patterns of workaround behaviour have been identified in other parts of the healthcare system. These studies highlight the potential threats to patient safety and quality of care, for example in the context of workarounds in medical systems (Koppel et al., 2008), while in other studies such practices are deemed necessary for healthcare practitioners to flourish and excel in a sociotechnical environment (see for example workarounds in nursing (Lalley & Malloch, 2010) or clinicians in general (Braithwaite et al., 2009)). While our study did not address these trade-offs specifically in the context of capacity planning, understanding the potential trade-offs between centralised and decentralised planning in future studies would help identify any existing threats to care quality and efficiency.

Finally, we reflect on the diverse skill base and organisational positions of “planners” in our studied hospital setting. A planner could mean anything from someone whose sole responsibility is to plan and schedule in an administrative function, to the surgical specialists. This means that the background, skillsets and experiences vary, but also the position of influence within the hospital. As shown in other studies, responsibilities for capacity planning are not uniform even in the same country (van der Ham et al., 2020). This can cause frustrations and uncertainties within the job position itself, especially among those with more administrative functions. One of our main challenges in our application of the CFIR framework was to identify differences between intervention beneficiaries/end-users, leaders and facilitators, as these roles are not clearly defined. Any future implementation of scheduling interventions would have to be mindful of these differences and the exact responsibilities of those who would adopt and champion the new intervention.

### What we learned regarding design inputs for the innovation (the proposed blueprint schedule)

Given the identified networks and informal communication leading to schedule adjustments, the learning point for designing a new schedule is that it needs to work within such a culture and expect these necessary adjustments. The system would have to be adaptable or flexible to allow for extreme cases, whether this is designed centrally or allowed to be designed further on in the process (and some of this uncertainty is already considered in the proposed model). This would also mean accounting for differences such as: in surgeon’s time for completing their tasks, the difference in children, patients and carers needing more comforting, comorbidities that affect anaesthesia and the impact of the surgery. These would be in addition to the existing considerations in the proposed model which already include different surgeries requiring different lengths of stay in the ward.

We also learned that any new schedule should clearly demonstrate its advantages, for example that it reduces the time they need to execute a task or increases the capacity instead of taking a lot of time and the result being relatively small. It is important that this is also shown in advance, which ensures staff would adopt the new approach. In this case, this can therefore be a shadow scheme or pilot and expand from there.

### What we learned about the use of CFIR for a proposed (not trialled) OR/MS innovation

To our knowledge, the explicit use of implementation science frameworks is not extensively used in OR/MS (Lamé et al., 2025). A study closely related to ours is that by (Visintin et al., 2017), whose aim was to identify the features that would make a master surgery schedule optimisation model effective and easy to implement, thereby identifying actions that would facilitate their introduction and use. They used an action research approach alongside their schedule design process.

Monks (2015), publishing in Implementation Science, has argued that OR/MS have much to contribute to Implementation Science, in three ways: by helping structure an implementation problem (i.e. enabling stakeholders to gain shared understanding of a complex issue in a healthcare setting), as a prospective evaluation tool (by modelling/appraising intervention efforts before implementing them), and for aiding larger strategic decisions (Monks, 2015). Monks argument is that OR/MS can offer health services research much valuable implementation lessons, i.e. “the use of OR to conduct implementation science upfront before any action to alter a care pathway or service has been taken”. Our study aimed to bring OR/MS and implementation science closer together but in a different way: using existing implementation science approaches to improve our (and therefore more likely adopt) OR/MS research outputs.

In our case, some critical reactions to the innovation arose from the fact that it is not a fully developed, trialled, evidence-based innovation, but bringing this draft schedule along allowed participants to ‘think along’ the need for improving capacity management. This partly shows the benefit of already using implementation principles even when only designing an intervention, as argued by Taylor and Kowalkowski who argue for using implementation science for pilot studies, to inform future randomised controlled trials in healthcare (Taylor & Kowalkowski, 2021). Furthermore, it highlights the importance of piloting and trialling something to develop a form of evidence base before introduced into its ultimate health system setting (participants noted the importance of demonstrating the relative advantage of the new schedule, if it were to be adopted). However, OR/MS outputs are not often trialled and evaluated in practice, or their uptake is left to industrial partners ((van Lent et al., 2012) and (Brailsford et al., 2016)), calling for more pilot or trial studies or at least the need for evaluating new OR/MS outputs.

Overall, however, CFIR was helpful to give us a framework to explore different implementation challenges across the various domains. While as researchers we could have left these as open interviews, or simply guessed the potential implementation challenges ourselves, the questions provided a scaffolding to guide the identification of barriers and facilitators for adopting this new schedule. It also helped elicit actual recommendations specific to the design of the schedule (as identified in the previous section). Specifically, it also highlights the importance of bringing stakeholders along even for capacity planning, which is not a typically considered innovation within health services research.

## 4. CONCLUSION

In this study, we directly addressed an implementation gap for operations research by using an implementation science framework (CFIR) to identify barriers and facilitators for adopting a mathematically optimised surgery blueprint schedule within a children’s hospital.

Our study resulted in three sets of findings with implications for future work. First, we identified the implementation barriers and facilitators for introducing a new scheduling approach in our study’s setting. Implementation facilitators included a strong motivation among staff to optimise schedules given resource constraints and work burdens, as well positive reactions to an objectively designed mathematical scheduling tool. Implementation barriers included a resistance to change among some staff, and the need for more evidence about the new schedule’s benefits before implementation. Most strikingly, we identified a strong culture of operational adjustments made to schedules in staff’s current working practices, which is a challenge to any optimised and centralised scheduling tool. Furthermore, we identified that the diversity in skills and influence of different staff who have the role of a ‘planner’ in a hospital setting can lead to frustrations and uncertainties about who could adopt and champion a new scheduling innovation. Secondly, we identified specific design inputs that would support the adoption of a new proposed scheduling approach, including the required adaptability to different surgeon’s needs and those of the patients/families, as well as the need to strongly communicate the benefits of the new schedule versus current practice.

Finally, we found that the CFIR was a helpful tool for identifying specific innovation adoption factors for a proposed surgery scheduling model. By applying CFIR ‘early’, in this case, in a proposed schedule before it is designed or even trialled for full implementation, we were able to also identify elements that can feed back into the design of the schedule in future and be mindful of the implementation challenges, which could also be fed back into the innovation design. We propose that the use of implementation science approaches, such as CFIR in our case, in future studies can be helpful in bridging the gap between theory and practice for outputs from operations research and management sciences.

## ACKNOWLEDGEMENTS

We would like to thank all the staff at the academic children’s hospital who took part in the study, and those who enabled us to conduct the work.

## DATA AVAILABILITY STATEMENT

All original interview audio recordings, transcripts and informed consent forms from the interviews are stored under the university’s secure SURFDRIVE system for a limited period. Due to these containing personal and sensitive information, they are not made available publicly in accordance with the ethical approval for this study.

## TABLE LIST

Table 2 List of interview participants included in the study

## FIGURE LIST

Figure 5 Different planning stages in the hospital

Figure 2. Sample of surgery blueprint schedule conceptualisation shown to show how patient groups are categorised according to surgery duration and length of stay

Figure 3 Surgery blueprint schedule conceptualisation (as presented to the interviewees). Each bar represents an operating room. The letters represent the scheduled surgery groups. The fading colours represent the uncertain surgery duration.

Figure 4 Overview of planning/scheduling staff interviewed, and relationship to tactical and operational planning functions (numbers correspond to interviewed participants in Table 1). Orange boxes demonstrate how these planning levels correspond with general planning in Figure 1. Blue box indicates the surgery blueprint schedule which is the ‘innovation’ introduced in this study.

## Supplementary material

### Appendix A: Interview protocol

1. To check whether I have explained the detailed OR schedule correctly, could you please explain it to me now?
2. What are your initial thoughts on the proposed more detailed OR schedule?
3. What do you think about the fact that the more detailed diagram was created using a mathematical model?
4. What are the differences between the proposed more detailed OR schedule and the current planning system at this hospital? What advantages or disadvantages does the more detailed OR schedule have compared to the current planning system?
5. We are now proposing this innovation to the scheduling system, but is there another method you would like to use to improve OR patient scheduling?
6. By whom and in what way do you think it will ultimately be decided whether the more detailed OR schedule will be (partly) implemented in this hospital?
7. If the participant schedules the patients we ask question (a), otherwise question (b).

i. How confident are you that you can divide the patients on the waiting list into the different patient groups? Why?
ii. How confident are you that planners can divide the patients on the waiting list into the different patient groups? Why?

*Now there are a number of questions to get a better idea of the daily affairs at this hospital, so that we can see how this would influence a possible implementation*.

8. How do you communicate about patient planning with colleagues within your team (colleagues, supervisors, people you manage, managers)? How would this affect a possible introduction of the more detailed OR schedule?
9. To what extent are new ideas embraced and used to make improvements within this hospital? Is the organization flexible? Does this differ per team/department?

i. Can you give a recent example?
10. How would the more detailed OR schedule fit into the current workflow? How does it interact with the current planning process? What would go right/wrong?
11. How would the more detailed OR schedule affect how patients’ needs are met? These can be both encouraging and hindering factors.
12. To what extent do you discuss the hospital’s planning system with colleagues from other departments at this hospital or other hospitals? Does this influence your opinion about the more detailed OR schedule?
13. We have now talked about many different aspects of the more detailed OR schedule and a possible implementation, but is there actually a great need for this more detailed OR schedule? May have already been answered at 2.

i. Why or not?
ii. Do others see a need to use this more detailed OR schedule?
14. **Only ask central planners: Is there policy from, for example, the hospital or legislation from the government that could influence the possible implementation of the more detailed OR schedule?**
15. *Only submit to OPO/TPO: If it were decided that this more detailed OR schedule would be used in the hospital. What steps do you think are necessary to move from this initial more detailed planning to full implementation? Who would advocate the necessary changes? Who would work on the development?*
16. Is there anything else you would like to say that we have not discussed?

### Appendix B: Constructs used from CFIR to guide the interview questions

**Table B1.**
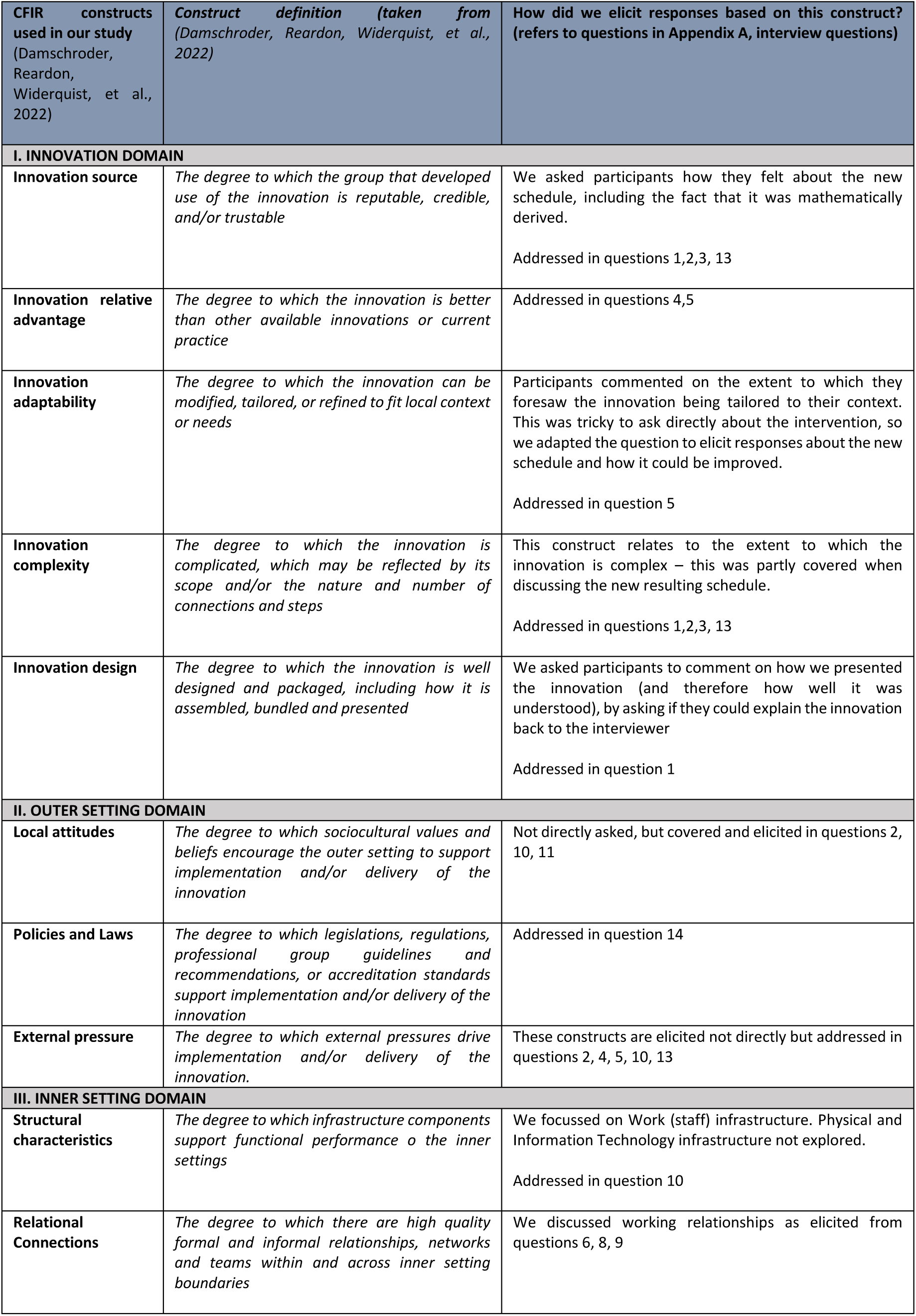
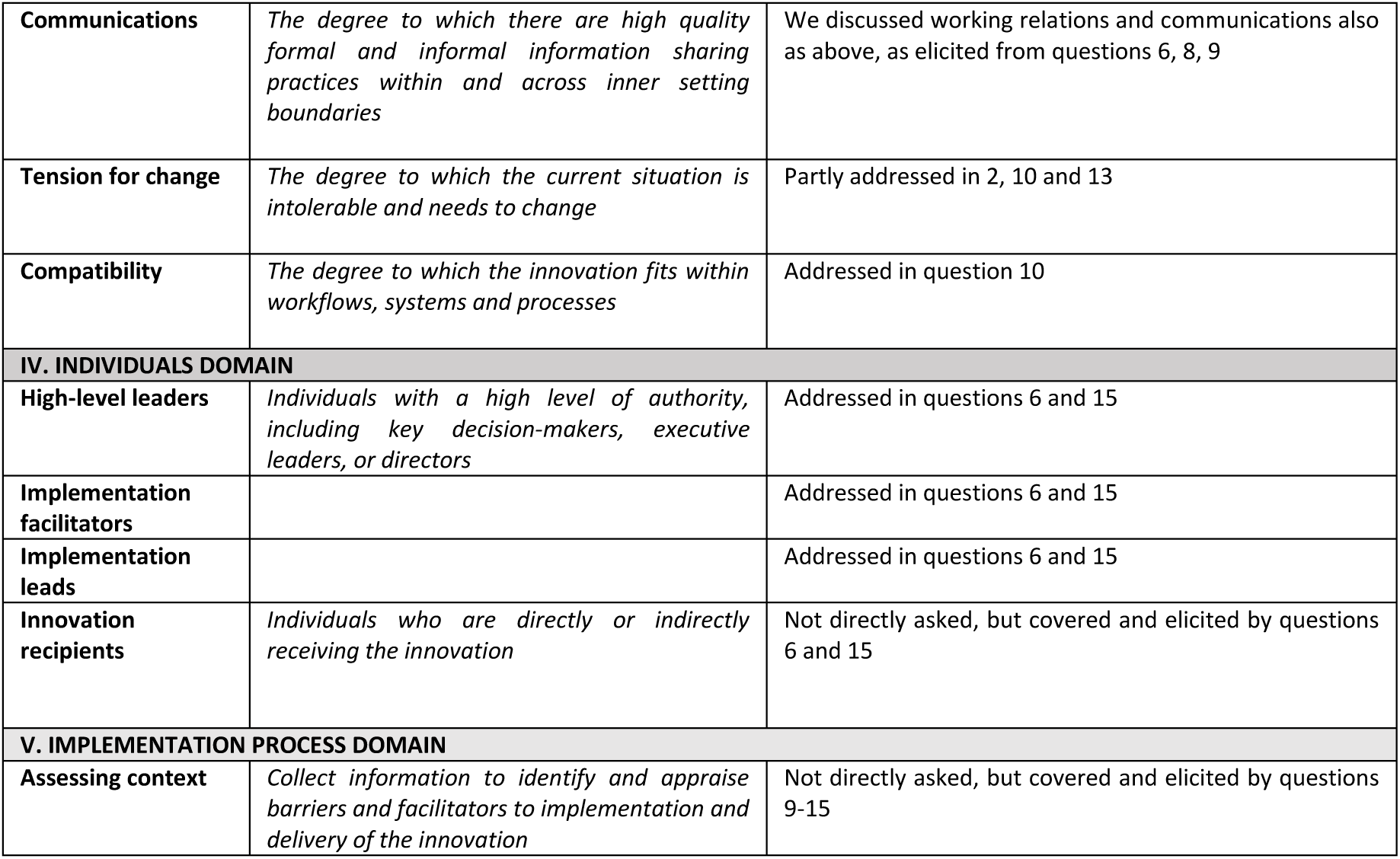
CFIR constructs explored in this study in relation to the new planning method (the “innovation”, according to (Damschroder, Reardon, Widerquist, et al., 2022))

| CFIR constructs used in our study (Damschroder, Reardon, Widerquist, et al., 2022) | Construct definition (taken from (Damschroder, Reardon, Widerquist, et al., 2022)) | How did we elicit responses based on this construct? (refers to questions in Appendix A, interview questions) |
| --- | --- | --- |
| **I. INNOVATION DOMAIN** |  |  |
| **Innovation source** | *The degree to which the group that developed use of the innovation is reputable, credible, and/or trustable* | We asked participants how they felt about the new schedule, including the fact that it was mathematically derived.<br>Addressed in questions 1,2,3, 13 |
| **Innovation relative advantage** | *The degree to which the innovation is better than other available innovations or current practice* | Addressed in questions 4,5 |
| **Innovation adaptability** | *The degree to which the innovation can be modified, tailored, or refined to fit local context or needs* | Participants commented on the extent to which they foresaw the innovation being tailored to their context. This was tricky to ask directly about the intervention, so we adapted the question to elicit responses about the new schedule and how it could be improved.<br>Addressed in question 5 |
| **Innovation complexity** | *The degree to which the innovation is complicated, which may be reflected by its scope and/or the nature and number of connections and steps* | This construct relates to the extent to which the innovation is complex – this was partly covered when discussing the new resulting schedule.<br>Addressed in questions 1,2,3, 13 |
| **Innovation design** | *The degree to which the innovation is well designed and packaged, including how it is assembled, bundled and presented* | We asked participants to comment on how we presented the innovation (and therefore how well it was understood), by asking if they could explain the innovation back to the interviewer<br>Addressed in question 1 |
| **II. OUTER SETTING DOMAIN** |  |  |
| **Local attitudes** | *The degree to which sociocultural values and beliefs encourage the outer setting to support implementation and/or delivery of the innovation* | Not directly asked, but covered and elicited in questions 2, 10, 11 |
| **Policies and Laws** | *The degree to which legislations, regulations, professional group guidelines and recommendations, or accreditation standards support implementation and/or delivery of the innovation* | Addressed in question 14 |
| **External pressure** | *The degree to which external pressures drive implementation and/or delivery of the innovation.* | These constructs are elicited not directly but addressed in questions 2, 4, 5, 10, 13 |
| **III. INNER SETTING DOMAIN** |  |  |
| **Structural characteristics** | *The degree to which infrastructure components support functional performance o the inner settings* | We focussed on Work (staff) infrastructure. Physical and Information Technology infrastructure not explored.<br>Addressed in question 10 |
| **Relational Connections** | *The degree to which there are high quality formal and informal relationships, networks and teams within and across inner setting boundaries* | We discussed working relationships as elicited from questions 6, 8, 9 |
| **Communications** | *The degree to which there are high quality formal and informal information sharing practices within and across inner setting boundaries* | We discussed working relations and communications also as above, as elicited from questions 6, 8, 9 |
| **Tension for change** | *The degree to which the current situation is intolerable and needs to change* | Partly addressed in 2, 10 and 13 |
| **Compatibility** | *The degree to which the innovation fits within workflows, systems and processes* | Addressed in question 10 |
| **IV. INDIVIDUALS DOMAIN** |  |  |
| **High-level leaders** | *Individuals with a high level of authority, including key decision-makers, executive leaders, or directors* | Addressed in questions 6 and 15 |
| **Implementation facilitators** |  | Addressed in questions 6 and 15 |
| **Implementation leads** |  | Addressed in questions 6 and 15 |
| **Innovation recipients** | *Individuals who are directly or indirectly receiving the innovation* | Not directly asked, but covered and elicited by questions 6 and 15 |
| **V. IMPLEMENTATION PROCESS DOMAIN** |  |  |
| **Assessing context** | *Collect information to identify and appraise barriers and facilitators to implementation and delivery of the innovation* | Not directly asked, but covered and elicited by questions 9-15 |

## Footnotes

1 This schedule was developed as part of an educational exercise for an MSc thesis. For details of its development, see: *[redacted for review]*

